# Convalescent plasma improves overall survival in patients with B-cell lymphoid malignancies and COVID-19: a longitudinal cohort and propensity score analysis

**DOI:** 10.1101/2021.12.29.21268525

**Authors:** Thomas Hueso, Anne-Sophie Godron, Emilie Lanoy, Jérôme Pacanowski, Laura I. Levi, Emmanuelle Gras, Laure Surgers, Amina Guemriche, Jean-Luc Meynard, France Pirenne, Salim Idri, Pierre Tiberghien, Pascal Morel, Caroline Besson, Rémy Duléry, Sylvain Lamure, Olivier Hermine, Amandine Gagneux-Brunon, Nathalie Freymond, Sophie Grabar, Karine Lacombe, the HEMOPLASM Study Group

## Abstract

Patients with hematological malignancies and COVID-19 display a high mortality rate. In such patients, immunosuppression due to underlying disease and previous specific treatment impair humoral response, limiting viral clearance. Thus, COVID-19 convalescent plasma (CCP) therapy appears as a promising approach through the transfer of neutralizing antibodies specific to SARS-CoV-2.

We report the effect of CCP in a cohort of 112 patients with hematological malignancies and COVID-19 and a propensity score analysis on subgroups of patients with B-cell lymphoid disease treated (n=81) or not (n=120) with CCP between 1 May 2020 and 1 April 2021. The overall survival of the whole cohort was 65% [56–74.9] and 77.5% [68.5–87.7] for patients with B-cell neoplasm. Prior anti-CD20 monoclonal antibodies therapy was associated with better overall survival whereas age, high blood pressure, and COVID-19 severity were associated with a poor outcome. After an inverse probability of treatment weighting approach, we observed in anti-CD20–exposed patients with B-cell lymphoid disease a decreased mortality of 63% (95% CI=31%–80%) in the CCP-treated group compared to the CCP-untreated subgroup, confirmed in the other sensitivity analyses.

Convalescent plasma may be beneficial in COVID-19 patients with B-cell neoplasm who are unable to mount a humoral immune response.

## Introduction

Patients with hematological malignancies and SARS-CoV-2 infection display a high mortality rate with an estimated risk of death of 34% that reaches 39% in hospitalized patients.^1,2^ In such patients, several studies highlighted that both underlying cellular or humoral immunosuppression may hamper virus clearance resulting in prolonged shedding and higher risk of severe COVID-19.^3,4^ Furthermore, anti-SARS-CoV-2 vaccine response in patients with hematological malignancies is lower compared to the general population, especially in patients with B–cell lymphoid disease.^5,6^ Thus, therapeutic approaches to inhibit viral replication and enhance viral clearance are mandatory in this specific population.

Early transfusion of high titer COVID-19 convalescent plasma (CCP) has emerged as a promising therapy to target SARS-CoV-2 and achieve clinical recovery.^7,8,9^ In France, CCP has been proposed in a national monitored access program, notably to hospitalized COVID-19 patients with underlying immunosuppression such as patients with hematological malignancy. While most randomized trials have not reported a benefit of CCP in a general population with COVID-19,^10^ we observed that B-cell depleted patients with protracted COVID-19 may benefit from CCP transfusion along with a decrease of all inflammatory parameters, oxygen weaning, and viral clearance.^11^ Accordingly, a retrospective propensity score matched analysis of 966 patients with a wide range of hematological malignancies, among whom 143 received CCP, and reported that CCP transfusion was associated with 40% lower mortality,^12^ without taking into consideration immortal time bias and specificity of each hematological malignancy.^13^ Building on these encouraging results, we report the outcome after CCP transfusion in a cohort of COVID-19 patients with hematological malignancies as well as on the results of a nested comparison of the survival among patients with B-cell neoplasm treated or not with CCP.

## Methods

### Patients and inclusion criteria

We analyzed all patients with hematological malignancies and virologically-documented COVID-19 included from 1 May 2020 to 1 April 2021 in a CCP monitored access program implemented in France (CCP cohort). Underlying disease included B-cell lymphoid neoplasm (such as diffuse B-cell lymphoma (DLBCL), chronic lymphoid leukemia (CLL), follicular lymphoma (FL), mantle cell lymphoma (MCL), marginal zone lymphoma (MZL), or B-acute lymphoblastic leukemia (B-ALL)) and plasma cell neoplasm requiring treatment and myeloid neoplasm (myelodysplastic syndrome or acute myeloid leukemia).

The subset of patients with B-lymphoid neoplasm in the CCP cohort was then compared in a propensity score analysis to a cohort of patients with similar disease who were not treated with CCP in French hospitals during the same successive COVID-19 outbreak periods (Figure 1). Both cohorts were treated as per standard of care for COVID-19. Patients gave their written informed consent for the retrospective data collection, and ethical clearance was obtained from the French Infectious Diseases Society (IRB number: 00011642).

**Figure 1.**
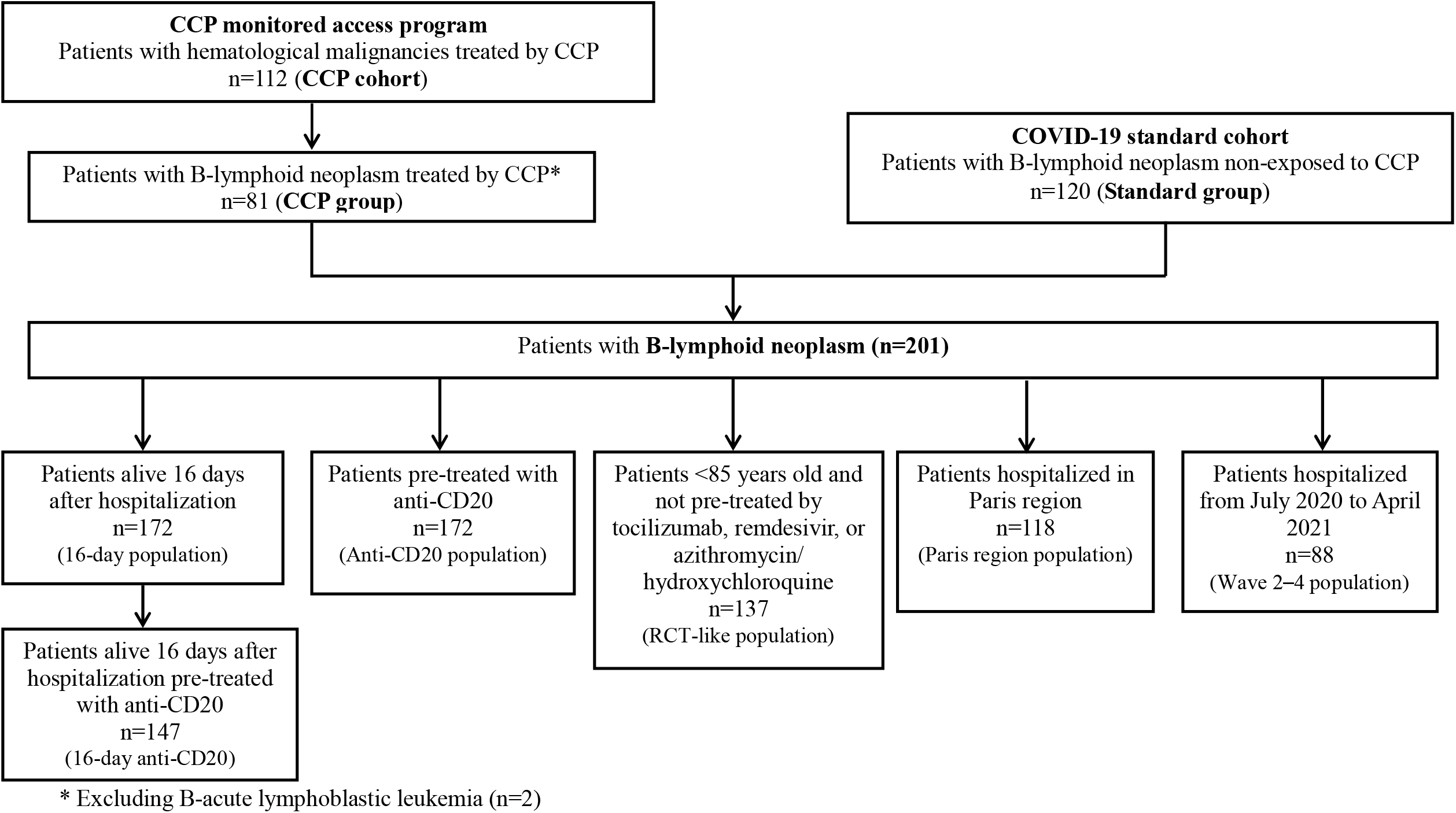
Flow chart

### Patient treatment and data collection

All patients included in the CCP monitored access program received four ABO-compatible CCP units (200–220 mL each), usually two units/day over two days (day 0 and day +1) and more rarely one unit/day over four days. Most often, patients received CCP from four different donors. Convalescent donors were eligible for plasma donation 15 days after resolution of COVID-19 symptoms. Collected apheresis plasma underwent pathogen reduction (Intercept blood system; Cerus, Concord, CA) and standard testing, as per current regulations in France. Additionally, anti–SARS-CoV-2 antibody content was assessed in each donation, with a requirement for a SARS-CoV-2 seroneutralization titer ≥40 (≥80 after October 2020) and/or an immunoglobulin G (IgG) enzyme-linked immunosorbent assay (EUROIMMUN, Bussy-Saint-Martin, France) ratio >5.6 (≥8 after October 2020).^14^

Clinical parameters (temperature and oxygen need) as well as grading on the WHO scale for COVID-19 severity and biological parameters, including inflammatory markers (C-reactive protein (CRP), ferritin, fibrinogen, and D-dimers), were recorded at the time of CCP transfusion (day 0).^15^ SARS-CoV-2 serological status and circulating lymphocyte subpopulations at day 0 were assessed. PCR in nasopharyngeal swab (with cycle threshold when available) was performed at day 0 and day +7 after CCP. Adverse events were recorded.

### Outcomes

The primary outcomes were 90-days overall survival and factors associated with the risk of death in all patients with hematological malignancies and COVID-19 after CCP transfusion treated in the monitoring access program. Then, among patients with B-cell neoplasm, overall survival from the first day of hospitalization for COVID-19 of patients who received CCP (CCP group) was compared to the survival of those who did not (standard group). Secondary outcomes included safety and kinetics of inflammatory parameters after CCP transfusion.

### Statistics

Continuous variables are described with their medians and interquartile ranges, whereas categorical variables are expressed as raw numbers and percentages. In the CCP cohort study, a Wilcoxon paired test was performed to compare clinical and biological parameters at day 0 and day +7 after CCP transfusion. Ninety-days overall survival (OS) was evaluated using Kaplan-Meier estimates from the time of CCP transfusion. A log-rank test was used to compare survival curves. Crude and adjusted hazards ratio (HR) of death were estimated by univariable and multivariable Cox proportional hazard model. The multivariable model was built after stepwise selection of the variables from the variables with p-value below 0.05 in univariable regressions. Covariates considered in univariable analysis were gender, age (≥70 years versus below), comorbidities (diabetes, high blood pressure, and body mass index), type of hematological malignancies, previous B-cell depletion therapy such as anti-CD20 or anti-CD19 monoclonal antibodies (mAbs), time between symptoms onset and CCP transfusion, and disease status (complete remission, partial remission/stable disease, and progressive disease).

To further evaluate the effectiveness of CCP in patients with B-cell neoplasms, we compared the 81 CCP-treated B-cell neoplasm patients (after exclusion of 2 patients with B-ALL) of the CCP cohort to 120 CCP-untreated B-cell neoplasm patients from the cohort partly described in Dulery et al. study (standard group).^16^ To estimate the effect of CCP on survival in non-randomized settings where patients’ characteristics associated with CCP exposure could differ in the two groups, an inverse probability of treatment weighting (IPTW) approach was retained in an effort to control for indication bias in several population analyses. An individual propensity score (i.e., the probability of treatment by CCP) was retrieved from a logistic regression model, including potential confounders, both associated with prognostic of survival and CCP indication: age (“<65 years” vs. “≥65 years”), comorbidity (high blood pressure or diabetes or BMI≥25kg/m^2^ yes vs. no), and corticosteroid therapy (yes vs. no). A pseudopopulation was built by weighting observations by propensity scores. In order to check if weighting improved the comparability of the two groups, absolute standardized differences were estimated in the unweighted and weighted samples. According to IPTW common practice, standardized differences <0.20 were considered negligible. For each analysis, the distribution of probability of receiving CCP in the two groups was examined visually in order to see if overlap existed and the positivity assumption was not violated.

Then, HRs of death associated with CCP were derived from weighted Cox proportional hazards. Estimates of HR of death associated with CCP from the univariable and multivariable (adjusted for variables included in the propensity score and described above) unweighted Cox proportional hazards model were also provided for the main and sensitivity analyses population. Of note, the number of variables included in both IPTW and multivariable models was limited by the relative low number of events; therefore, only variables strongly associated with indication of CCP and with sufficient numbers of patients for each modality were kept. Main analysis was restricted to patients with B-cell lymphoid disease previously treated with anti-CD20 therapy and who were alive 16 days after hospitalization for COVID-19, which corresponded to the median time of CCP transfusion after hospitalization. In order to limit the immortal time bias that can arise because patients treated with CCP had to survive long enough after hospitalization to receive CCP,^13^ a landmark approach was chosen. Follow-up of patients who received CCP after more than 16 days following hospitalization was censored at time of CCP initiation. Since the use of CCP could differ in terms of area and period of recruitment, and exposure to cancer therapies and to COVID-19 medications that were not accounted for in IPTW, several sensitivity analyses in which baseline was date of hospitalization were conducted to assess for the robustness of the main results: (i) in the overall population of patients exposed and non-exposed to CCP; (ii) in a population restricted to anti-CD20 therapy pre-treated patients, excluding patients over 85 years, and patients pre-exposed to tocilizumab, remdesivir, or azithromycin/hydroxychloroquine; (iii) in a population excluding the first epidemic wave from March 2020 to June 2020; and (iv) in a population restricted to Paris region (Ile-de-France area, see Figure 1 and Table S1). Another sensitivity landmark analysis with baseline at date of hospitalization plus 16 days has been carried out in patients alive 16 days after hospitalization. Of note, other sensitivity analyses on overall population were also adjusted for gender. All analyses were done using R software version 3.6.1 and SAS® Software version 9.4 (Cary, North Carolina, USA). A p-value below 0.05 denoted statistical significance.

## Results

### COVID-19 convalescent plasma cohort analysis

One hundred and twelve patients (33 F/79 M) aged 62.5 (range) (20−88) years with hematological malignancies and COVID-19 received CCP. Among them, 83 (74%) patients were treated for B-lymphoid neoplasm, 10 (9%) for myeloid neoplasm, and 19 (17%) for multiple myeloma. Eighty-one (72%) patients had received anti-CD20 or anti-CD19 targeted therapy at a median of 42 (137−14) days before first symptoms of COVID-19. Ninety-eight patients (87%) patients had a negative SARS-CoV-2 serology at the time of CCP transfusion. The median circulating B-lymphocytes count was 0 (interquartile range) (0−0)/mm^3^ in 58 evaluable patients with B-cell lymphoid disease, 9 (0−48)/mm^3^ in 6 patients with myeloid neoplasm, and 7 (1−–17)/mm^3^ in 9 patients with plasma cell neoplasm. Of note, 11/83 (13%) patients with B-lymphoid neoplasm, 2/10 (20%) with myeloid neoplasm, and 8/19 (42%) with plasma cell neoplasm were mechanically ventilated (WHO scale 7), at the time of CCP transfusion. Previous COVID-19 treatments included corticosteroids (n=72, 64%), tocilizumab (n=8, 7%), and remdesivir (n=13, 12%). No patients received anti-spike mAbs or were vaccinated. The remaining patient characteristics are described in Table 1.

**Table 1.** Convalescent Plasma therapy (CPP cohort) characteristics

| **Patient characteristics** | **Patients**<br>**n=112** |
| --- | --- |
| **Age, mean (range)** | 62.5 (20–88) |
| **B-lymphoid neoplasm, n (%)** | 83 (74%) |
| Diffuse large B-cell lymphoma | 28 (25) |
| Follicular lymphoma | 23 (20) |
| Mantle cell lymphoma | 11 (10) |
| Marginal zone lymphoma | 4 (3) |
| Chronic lymphoid leukemia | 13 (12) |
| Waldenström Macroglobulinemia | 2 (2) |
| B-acute lymphoblastic leukemia | 2 (2) |
| **Myeloid neoplasm, n (%)** | 10 (9%) |
| Myelodysplastic syndrome | 2 (2) |
| Acute myeloid leukemia | 8 (7) |
| **Plasma cell neoplasm, n (%)** | 19 (17) |
| **Previous anti-CD19 or anti-CD20 mAbs, n (%)** | 81 (72) |
| **Previous anti-CD38 mAbs, n (%)** | 10 (9) |
| **Median time between last anti-CD19 or anti-CD20 mAbs**<br>**and COVID-19, days (IQR)** | 42 (137–14) |
| **Previous stem cell transplantation, n (%)** |  |
| Autologous | 27 (25) |
| Allogeneic | 5 (4) |
| **Disease status at the time of COVID-19, n (%)** |  |
| Complete remission | 48 (43) |
| Partial remission/stable disease | 29 (25) |
| Progressive disease | 22 (20) |
| Missing data | 13 (12) |
| **B-lymphocyte count at the time of CCP transfusion,**<br>**median (IQR)** |  |
| B-lymphoid neoplasm | 0 (0–0) |
| Myeloid neoplasm | 9 (0–48) |
| Plasma cell neoplasm | 7 (1–17) |
| **Serological status for COVID-19, n (%)** |  |
| Positive | 2 (2) |
| Negative | 98 (87) |
| Unknown | 12 (11) |
| **Previous COVID-19 specific treatment, n (%)\*** |  |
| Steroid | 72 (64) |
| Tocilizumab | 8 (7) |
| Remdesivir | 13 (12) |
| Others | 8 (7) |
| Nothing | 32 (29) |

\* Some patients may have simultaneous treatment; mAbs: Monoclonal antibodies; IQR: Interquartile range.

Transfusion-related adverse events were reported in a limited number of patients and included 3 allergies (2 minor, 1 severe) and 2 cases of transient increase in oxygen requirement of uncertain imputability. CCP transfusion was associated with decreased (d+7 vs. d0) temperature (p<0.0001), CRP (p<0.0001), ferritin (p<0.0001), and fibrinogen (p=0.026). Conversely, PCR cycle threshold values significantly increased (p=0.00015) (Figure 2). The 90-days overall survival of whole cohort was 64.9% (95% CI=56.2−3;74.9), 77.5% (95% CI=68.5−87.7) in patients with B-lymphoid neoplasm, 20% (95% CI=5.8−69.1) with myeloid neoplasm, and 36.8% (95% CI=20.4−66.4) with plasma-cell neoplasm. Deaths were all associated with COVID-19. In univariable analysis, age >70 years (p=0.006), high blood pressure (p=0.003), the type of hematological disease (p<0.0001), a previous anti-CD20/CD19 mAbs (p<0.0001), COVID-19 severity (p=0.0004), and the time of CCP administration (p=0.002) were significantly associated with overall survival (Table 2 and Figure 3). In multivariable analysis, only age >70 yrs (HR=2.63, 95% CI=1.31−5.27; p=0.008), high blood pressure (HR=2.71, 95% CI=1.22−6.06; p=0.015), and COVID-19 severity (WHO scale 6 & 7) (HR=5.36, 95% CI=1.12−25.64 and HR=6.31, 95% CI=1.31−30.39; p=0.038 respectively) were associated with lower overall survival, whereas previous anti-CD20/CD19 mAbs was strongly associated with better overall survival (HR=0.22, 95% CI=0.10−0.51; p<0.001) (Figure 4).

**Figure 2:**
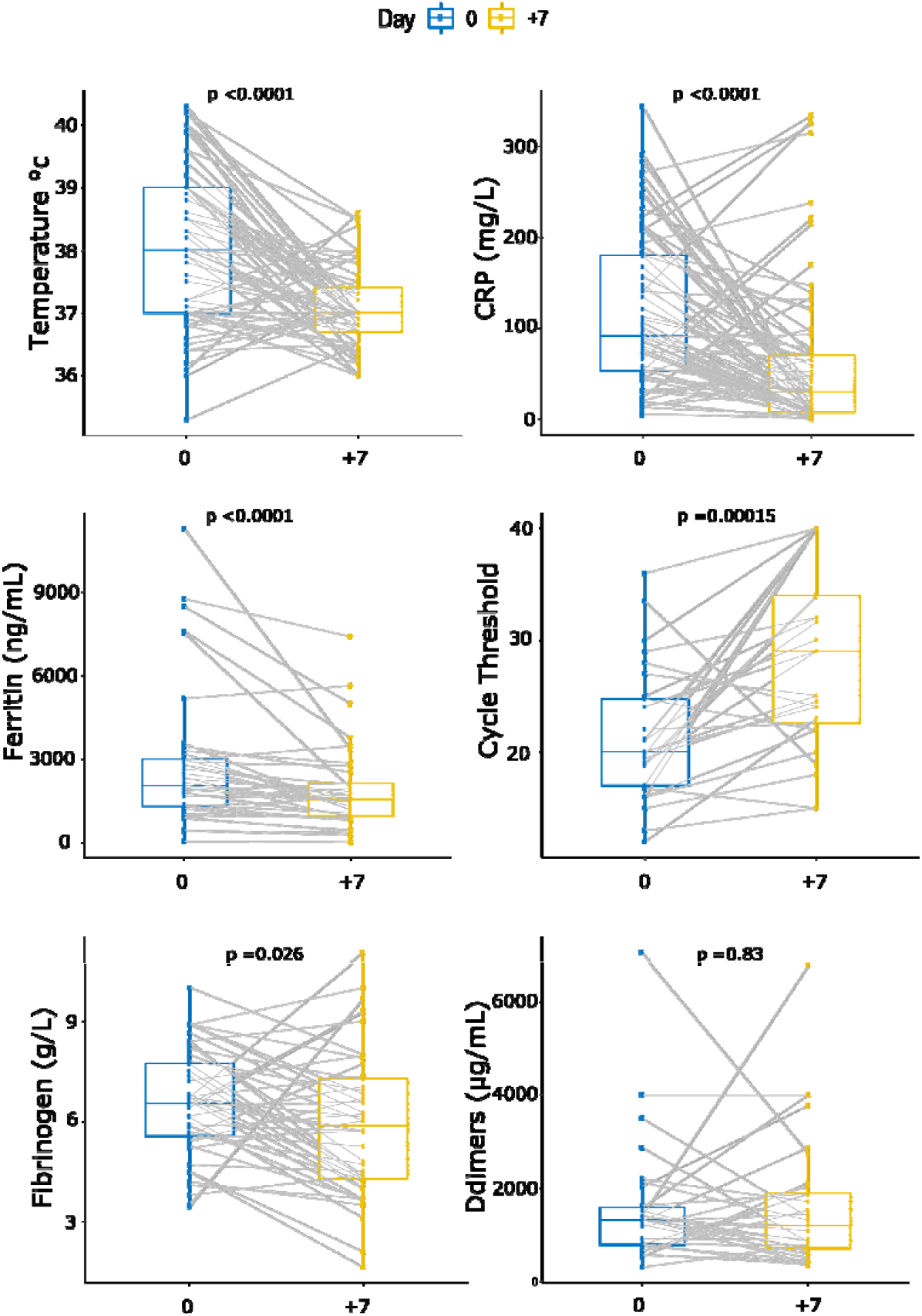
Kinetics of inflammatory parameters. All parameters were assessed the day of convalescent plasma transfusion (day 0) and 7 days after (day +7). A cycle threshold value over 40 was considered negative. A Wilcoxon-paired test was assessed. The median and interquartile ranges are represented.

**Table 2.**
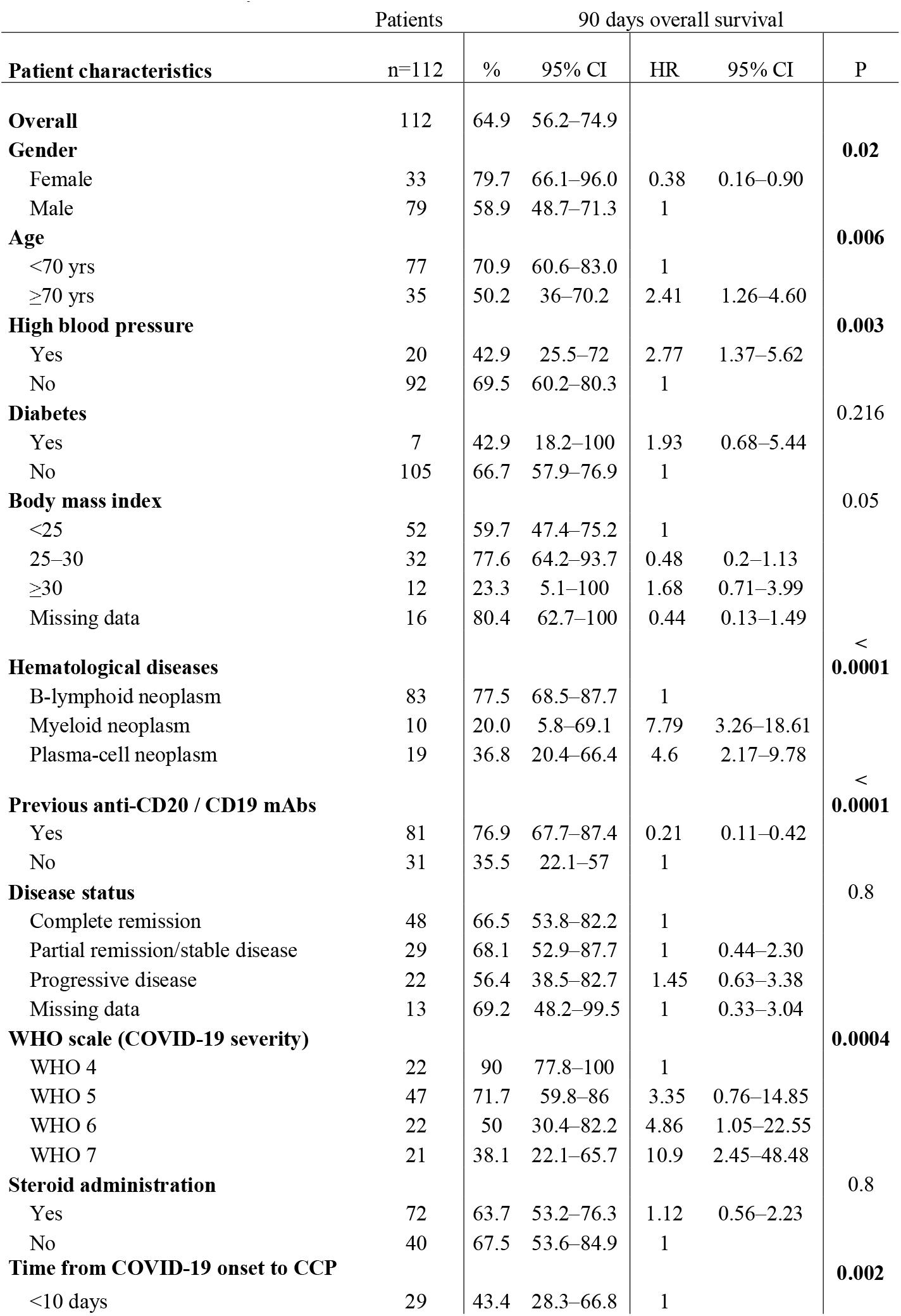
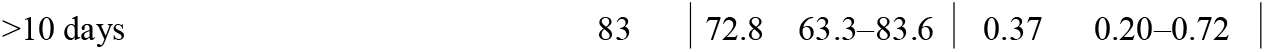
Univariable analysis

**Table 3.** Characteristics of patients with B-lymphoid neoplasm according to analyzed population (main analysis, overall population analyses).

|  | Analyzed population |  |  |  |
| --- | --- | --- | --- | --- |
|  | 16d anti-CD20 population |  | Overall population |  |
|  | Group |  | Group |  |
|  | CP+ | CP- | CP+ | CP- |
|  | N (%) | N (%) | N (%) | N (%) |
| **All** | **32** | **115** | **81** | **120** |
|  | (100%) | (100%) | (100%) | (100%) |
| **Epidemic wave** |  |  |  |  |
| 1st - Mar–Jun 2020 | 2 (6%) | 81 (71%) | 19 (23%) | 94 (77%) |
| 2nd - Jul–Oct 2020 | 8 (25%) | 20 (17%) | 26 (32%) | 10 (8%) |
| 3rd - Nov–Dec 2020 | 18 (56%) | 13 (11%) | 32 (40%) | 11 (9%) |
| 4th - Jan–Apr 2021 | 4 (13%) | 1 (1%) | 4 (5%) | 5 (4%) |
| **Geographic area** |  |  |  |  |
| Paris region | 13 (41%) | 69 (60%) | 33 (41%) | 85 (71%) |
| Eastern France | 1 (3%) | 26 (23%) | 9 (11%) | 30 (25%) |
| Other | 18 (56%) | 20 (17%) | 39 (48%) | 5 (4%) |
| **Gender** |  |  |  |  |
| Male | 19 (59%) | 76 (66%) | 56 (69%) | 72 (60%) |
| Female | 13 (41%) | 39 (34%) | 25 (31%) | 48 (40%) |
| **Age (years)** |  |  |  |  |
| <55 | 9 (28%) | 17 (15%) | 18 (22%) | 11 (9%) |
| 55–64 | 11 (35%) | 41 (36%) | 27 (33%) | 35 (29%) |
| 65–74 | 9 (28%) | 37 (32%) | 24 (30%) | 36 (30%) |
| ≥75 | 3 (9%) | 20 (17%) | 12 (15%) | 38 (32%) |
| **BMI (kg/m<sup>2</sup>)** |  |  |  |  |
| <18.5 | 2 (6%) | 5 (4%) | 3 (4%) | 6 (5%) |
| [18.5;25[ | 14 (44%) | 56 (49%) | 34 (41%) | 61 (51%) |
| [25;30[ | 10 (31%) | 29 (25%) | 24 (30%) | 29 (24%) |
| ≥30 | 4 (13%) | 15 (13%) | 8 (10%) | 17 (14%) |
| Missing | 2 (6%) | 10 (9%) | 12 (15%) | 7 (6%) |
| **Arterial hypertension** |  |  |  |  |
| No | 27 (84%) | 80 (70%) | 69 (85%) | 70 (58%) |
| Yes | 5 (16%) | 35 (30%) | 12 (15%) | 50 (42%) |
| **Diabetes** |  |  |  |  |
| No | 30 (94%) | 98 (85%) | 76 (94%) | 93 (77%) |
| Yes | 2 (6%) | 17 (15%) | 5 (6%) | 27 (23%) |
| **AH or diabetes or BMI >25kg/m<sup>2</sup> - PS\*** |  |  |  |  |
| No | 14 (44%) | 52 (45%) | 41 (51%) | 45 (38%) |
| Yes | 18 (56%) | 63 (55%) | 40 (49%) | 75 (63%) |
| **WHO scale at hospitalization** |  |  |  |  |
| Missing | 0 (0%) | 2 (2%) | 0 (0%) | 6 (5%) |
| WHO 4 | 8 (25%) | 41 (36%) | 29 (36%) | 33 (28%) |
| WHO 5 | 22 (69%) | 58 (50%) | 44 (55%) | 69 (57%) |
| WHO 6 | 2 (6%) | 7 (6%) | 6 (7%) | 6 (5%) |
| WHO 7 | 0 (0%) | 7 (6%) | 2 (2%) | 6 (5%) |
| **Anti-CD20 mAbs** |  |  |  |  |
| No | 0 (0%) | 0 (0%) | 3 (4%) | 26 (22%) |
| Yes | 32 (100%) | 115 (100%) | 78 (96%) | 94 (78%) |
| **Corticotherapy - PS\*** |  |  |  |  |
| No | 11 (34%) | 67 (58%) | 29 (36%) | 83 (69%) |
| Yes | 21 (66%) | 48 (42%) | 52 (64%) | 37 (31%) |
| **Tocilizumab** |  |  |  |  |
| No | 31 (97%) | 106 (92%) | 75 (93%) | 115 (96%) |
| Yes | 1 (3%) | 9 (8%) | 6 (7%) | 5 (4%) |
| **Azithromycin-hydroxychloroquine** |  |  |  |  |
| No | 32 (100%) | 107 (93%) | 79 (98%) | 109 (91%) |
| Yes | 0 (0%) | 8 (7%) | 2 (2%) | 11 (9%) |
| **Remdesivir** |  |  |  |  |
| No | 29 (91%) | 107 (93%) | 72 (89%) | 117 (98%) |
| Yes | 3 (9%) | 8 (7%) | 9 (11%) | 3 (3%) |
| **B-cell lymphoid neoplasm** |  |  |  |  |
| Chronic lymphoid leukemia | 6 (19%) | 11 (10%) | 13 (16%) | 18 (15%) |
| Diffuse large B-cell lymphoma | 12 (37%) | 48 (41%) | 28 (35%) | 49 (42%) |
| Follicular lymphoma | 6 (19%) | 32 (28%) | 23 (28%) | 22 (18%) |
| Marginal zone lymphoma | 1 (3%) | 10 (9%) | 4 (5%) | 16 (13%) |
| Mantle cell lymphoma | 5 (16%) | 13 (11%) | 11 (14%) | 11 (9%) |
| Waldenström Macrglobulinemia | 2 (6%) | 1 (1%) | 2 (2%) | 4 (3%) |

The proportion of treated patients differed greatly among the strata of variables included in the propensity score such as age: 54% of treated in <65 years vs. 36% in ≥65 years in RCT-like population, 48% of treated in absence of comorbidity vs. 35% in presence of comorbidity in overall population, 14% of treated in patients not receiving corticotherapy vs. 30% in patients receiving corticotherapy. Moreover, this table shows that the proportion of treated patients differed also for variables that could not be included in the propensity score but defined sensitivity analysis populations such as wave, pre-exposure to anti-CD20, and area of hospitalization. AH: arterial hypertension; BMI: body mass index; PS\*: included in the propensity score; N: number of patients; WHO: World Health Organization; mAbs: monoclonal antibodies

**Figure 3:**
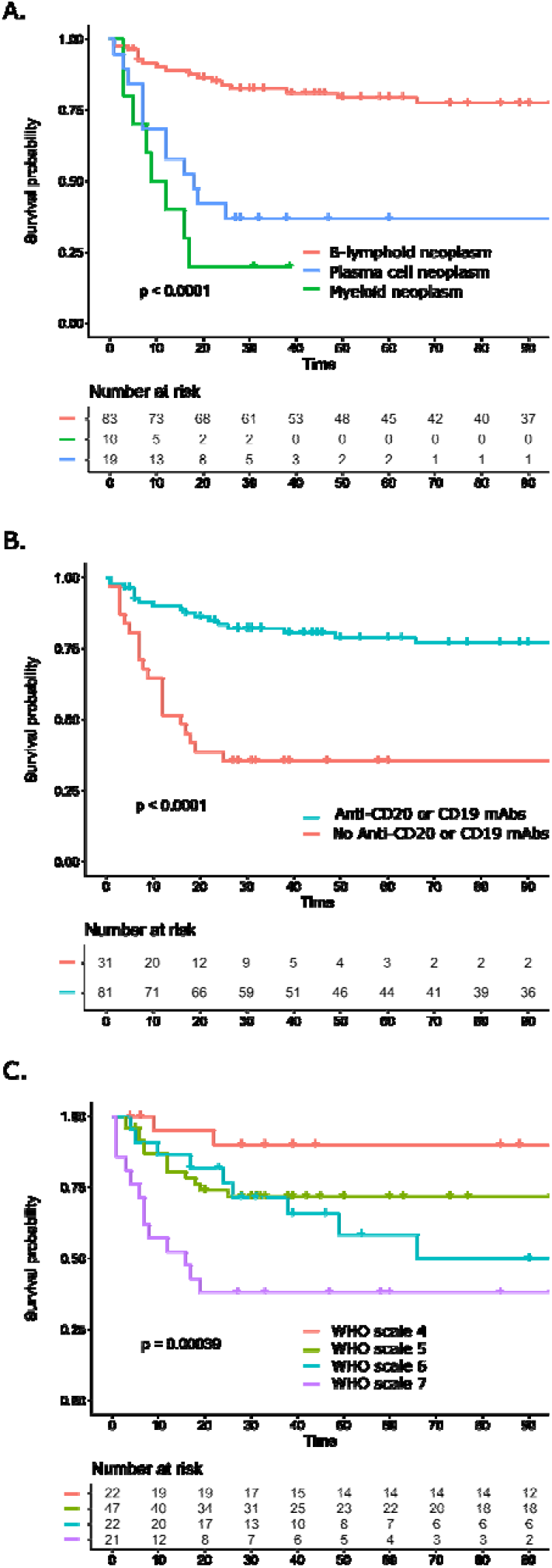
Overall survival after COVID-19 convalescent plasma transfusion represented using the Kaplan-Meier method according to hematological malignancies (A); the exposition to anti-CD20 or anti-CD19 monoclonal antibodies (mAbs) (B); and the COVID-19 severity (WHO scale) (C).

**Figure 4:**
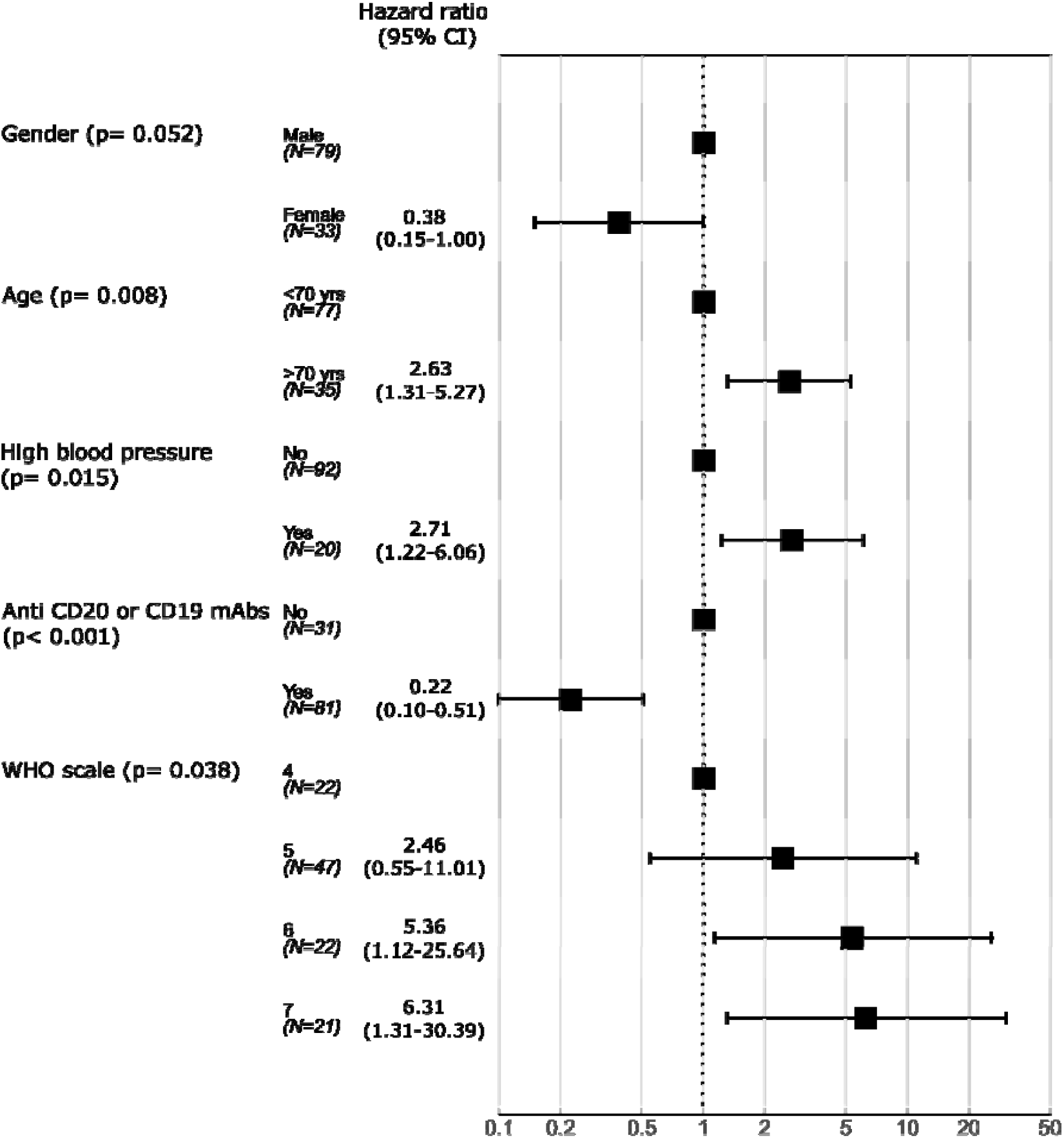
Forest plot representing hazard ratio (HR) of death obtained in multivariable analysis. mAbs: monoclonal antibodies

### Propensity score analysis in patients with B-lymphoid neoplasm

The numbers of patients included in each population analysis are provided in the flow chart in Figure 1. The main population analysis included 147 patients pre-exposed to anti-CD20 therapy and alive on day 16 after hospitalization for COVID.

The characteristics of patients of each analyzed population are detailed in Supplementary Table S1. This table shows that patients treated with CCP differ from the untreated ones, including for variables that could not be included in the propensity score. To account for the latter variables, sensitivity analysis populations restricted to specific wave, to pre-exposure to anti-CD20, or to specific area of hospitalization were performed.

In the main IPTW analysis, the exposure to CCP was associated with a 63% (95% CI=31%−80%) decrease in the risk of death in the 16-day anti-CD20 pre-exposed population, The HR estimated by IPTW varied across the different analysis populations from 40% in the waves 2– 4 population to 63% in the main analysis population as shown in figure 5. Multivariable models provided similar results to IPTW (Supplementary table S2). The addition of gender to the propensity score did not modify the estimation of the effect of the convalescent plasma exposure. Hazard ratios of death associated with each variable included in the propensity score are provided in Supplementary Table S3.

**Figure 5.**
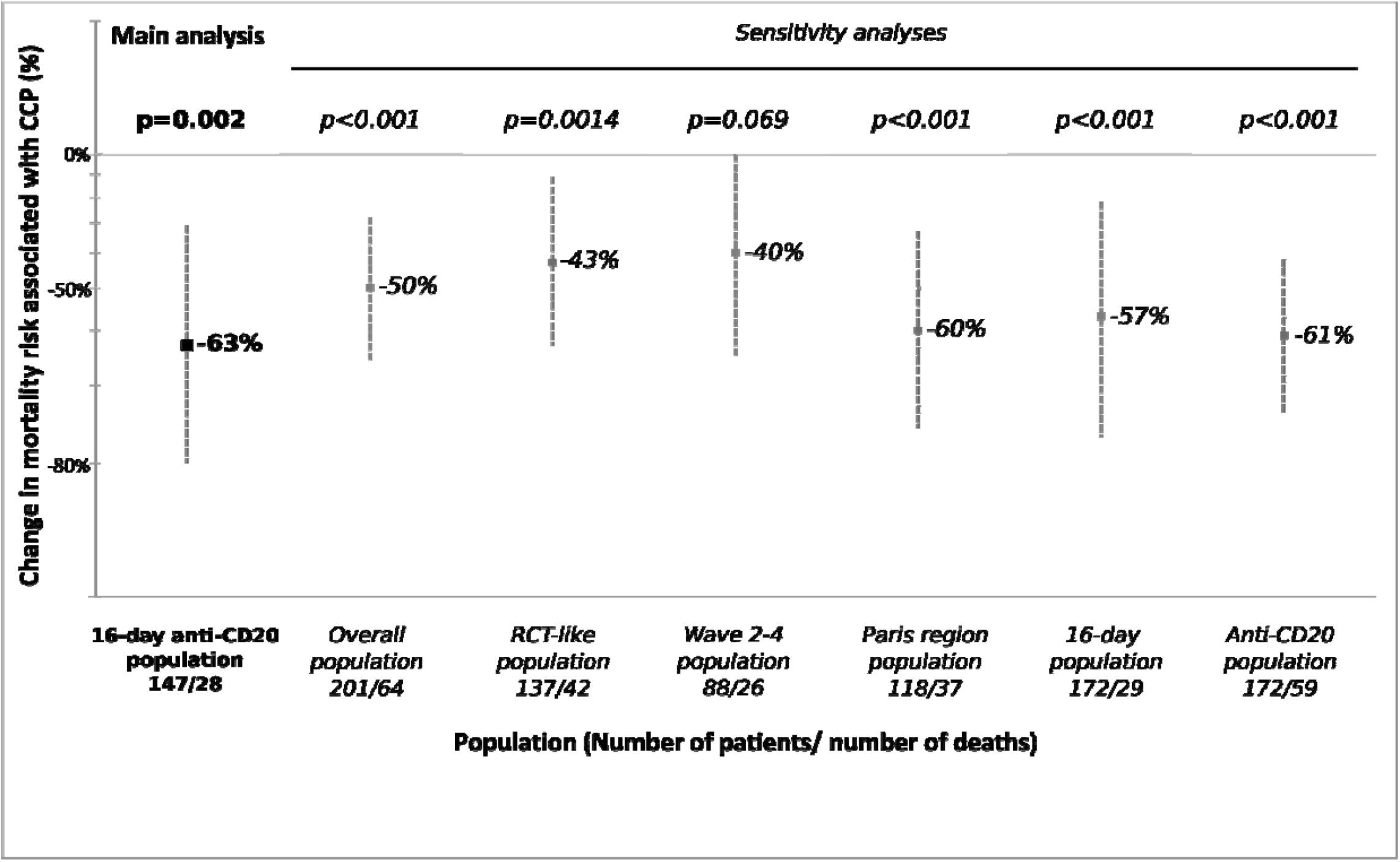
Change (dot) and its 95% confidence interval (line) in mortality risk associated with Covid19 convalescent plasma (CCP) therapy from inverse-probability weighting (IPTW) model

## Discussion

Our study reports the effect of CCP in a large and representative cohort of patients with hematological malignancies and who were hospitalized for severe COVID-19. Overall, 90-days survival was 64.9% for the whole cohort and 77.9% in the B-cell neoplasm subgroup with an overall survival benefit estimated between 43 % and 63 % after IPTW approach supporting the results of the Thompson et al. study.^12^ We also report a significant decrease of all inflammatory parameters as well as negativation of the nasopharyngueal PCR swab at d+7, consistent with our previous report.^11^ Furthermore, CCP infusion was well tolerated.^17^ As previously described, age and comorbidities such as high blood pressure and COVID-19 severity emerged as the most important risk factors of death due to COVID-19.^18^ Interestingly, although an increased risk of death was associated with B-depletion therapy such as anti-CD20 or anti-CD19 mAbs in a recently published retrospective lymphoma cohort,^16^ previous B-cell therapy depletion was strongly associated with better overall survival in patients receiving convalescent plasma even after adjustment for confounding factors. Since most patients in our study (87%) transfused with CCP had a negative serology for COVID-19, we assume that such patients were unable to produce neutralizing SARS-CoV-2 antibodies due to the underlying hematological disease or the previous administered drugs targeting B-cells.^19^ Subsequent transfer of neutralizing antibodies in such patients by means of CCP transfusion resulted in the control of viral replication and allowed for clinical recovery. The poor benefit of CCP transfusion in patients with plasma cell or myeloid neoplasm as well as the uncertain benefit of CCP in the general COVID-19 population able to mount humoral response supports our hypothesis.

To further investigate the effect of CCP, we compared the subset of CCP-treated patients with B-cell lymphoid malignancies and COVID-19 to a similar cohort of patients who were treated with standard of care only. The methodology sought to control for biases inherent to the observational design of this retrospective comparison; this included indication and immortal time biases. We showed that patients with B-cell lymphoid disease and pre-exposed to anti-CD20 therapy treated with CCP exhibited a significantly better survival probability than those who did not receive CCP. This important finding supports the results reported in a US-retrospective cohort^12^ as well as the positive trend observed in immunosuppressed patients (odds ratio (OR) 1.51 (0.80 to 2.92)) included in the recently reported REMAP-CAP trial^20^ and confirms the essential role of convalescent plasma in the COVID-19 therapeutics landscape of patients with B-cell malignancies.

However, those results present several limitations and raise several questions. In our longitudinal cohort, the risk of death reaches 80% and 64% for myeloid neoplasm and plasma cell neoplasm, respectively, that is twofold higher than reported in previous studies.^1^ We must point out a higher COVID-19 severity with 34% of patients with myeloid and plasma-cell neoplasm requiring for mechanical ventilation whereas only 13% for B-cell neoplasm at the time of CCP. Similarly, previous studies reported a better outcome after early CCP transfusion in non-hospitalized patients.^21^ Of note, we observed a lower overall survival in patients transfused within the first ten days after symptoms onset in univariable analysis, an association that disappeared in the multivariable analysis. A similar observation was made in the Recovery trial (in the CCP arm as well as the control arm) and may reflect more severe disease in patients hospitalized early in the course of the disease.^22^ Furthermore, 62% of patients transfused earlier (within the first ten days) transfusion in our cohort presented with myeloid or plasma-cell neoplasm.

In our retrospective nested analysis, data were collected prospectively from two data sources: one for patients exposed to CCP and another one for patients not exposed to CCP. Such comparison could not identify causal effect of treatment exposure with the level of proof provided by randomized clinical trials. We have tried to limit bias by using statistical methods to control indication bias (propensity score) and the immortal bias (time-lead analysis). The benefit of CCP appears to be robustly identified in all our sensitivity analysis. In addition, the main analysis, which allows for stricter controls on indication and immortal bias (use of 16-day landmark and IPTW identifies the strongest effect. We also have no data on the type of viral strain that infected the patients who received CCP, and thus could not analyze the impact of virological parameters in the response to CCP. However, during the inclusion period, SARS-CoV-2 strains circulating in France were mainly the Wuhan original strain and its Alpha variant. Therefore, we cannot extend our results to the Delta variant of concern that is currently becoming dominant worldwide. Plasma from convalescent and vaccinated donors exhibit high titer crossvariant antibodies that may provide a higher efficacy than the CCP used in our study.^23,24,25^ We presently favor the issuing of such CCP for treatment of immunosuppressed patients with B-cell malignancies.^26^

Although our study generates some strong arguments in favor of the use of CCP in B-cell depleted COVID-19 patients, several recent issues need to be considered. Indeed, initiation of the anti-SARS-CoV-2 vaccination,^27^ the availability and use of anti-spike mAbs in patients at risk of developing a severe form of COVID-19,^28^ and the emergence of SARS-CoV-2 variants have changed the course of COVID-19 outbreak. Firstly, none of the patients in both cohorts (CCP-treated and non-CCP-treated) were vaccinated against SARS-CoV-2. Although prospective randomized trials strongly support the efficacy of vaccination in the general population, results are less favorable in immunosuppressed patients, especially patients with CLL or multiple myeloma who exhibit a lower serological response despite two administrations of BNT162b2 mRNA COVID-19 vaccine.^5,29,30^ Further investigations are needed to evaluate the efficacy of vaccination on mortality linked to COVID-19 in such patients. Secondly, use of anti-spike mAb opened a new perspective, especially in frail patients with a reduction of related hospitalization or death in mild to moderate COVID-19 patients.^31^ Besides the cost of such an approach (over 2000 euros per administration compared to ≈120 euros/CCP) and possibly limited availability, immune escape mutations have been described especially in patients with B-cell lymphoid disease, justifying a successful treatment rescue with CCP.^32,33^ In light of this, emergence of variants due to protracted shedding or therapy-related selection^34,35^ remains the most important challenge to face. In our series, although we did not assess the SARS-CoV-2 lineages, we did not observe a difference of overall survival between the three epidemic outbreaks.

In conclusion, among patients with hematological malignancies and COVID-19, CCP represents an interesting approach, especially in patients with B-cell lymphoid malignancies and who are pre-treated with anti-CD20 mAbs. Importantly, such patients have poor response to vaccination and may present an escape variant due to prolonged shedding or anti-spike mAbs administration. The role of CCP in the treatment of COVID-19 patients unable to mount a humoral response could be strengthened if confirmed in a randomized prospective trial and should also be discussed in light of the increasing availability of anti-spike mAbs.

## Data Availability

All data produced in the present study are available upon reasonable request to the authors

## Author contribution

T.H., K.L., P.T. C.B. and R.D. designed the study and wrote the manuscript. S.G. and E.L. analyzed data and performed statistical analyses. T.H., AS.G., J.P., JL.M., LI.L., E.G., L.S., A.G., F.P., S.I., P.M., S.L., O.H., A.GB., N.F managed and followed-up patients, collected data, revised and approved manuscript.

## Acknowledgements

The authors would like to acknowledge all the clinicians for their participation in data collection. We also thank Dorothée Fey, who managed the weekly multidisciplinary convalescent plasma review meetings and participated in the data collection; the CCP donors; Etablissement Français du Sang personnel who managed CCP collection and testing; as well as the patients who consented to the anonymous use of their clinical data for this study.

This study was supported by the program Emergency Support Instrument (ESI) of the European Commission (project CCPEFS, grant number 101021793).

## Supplementary Tables

**Table S1.**
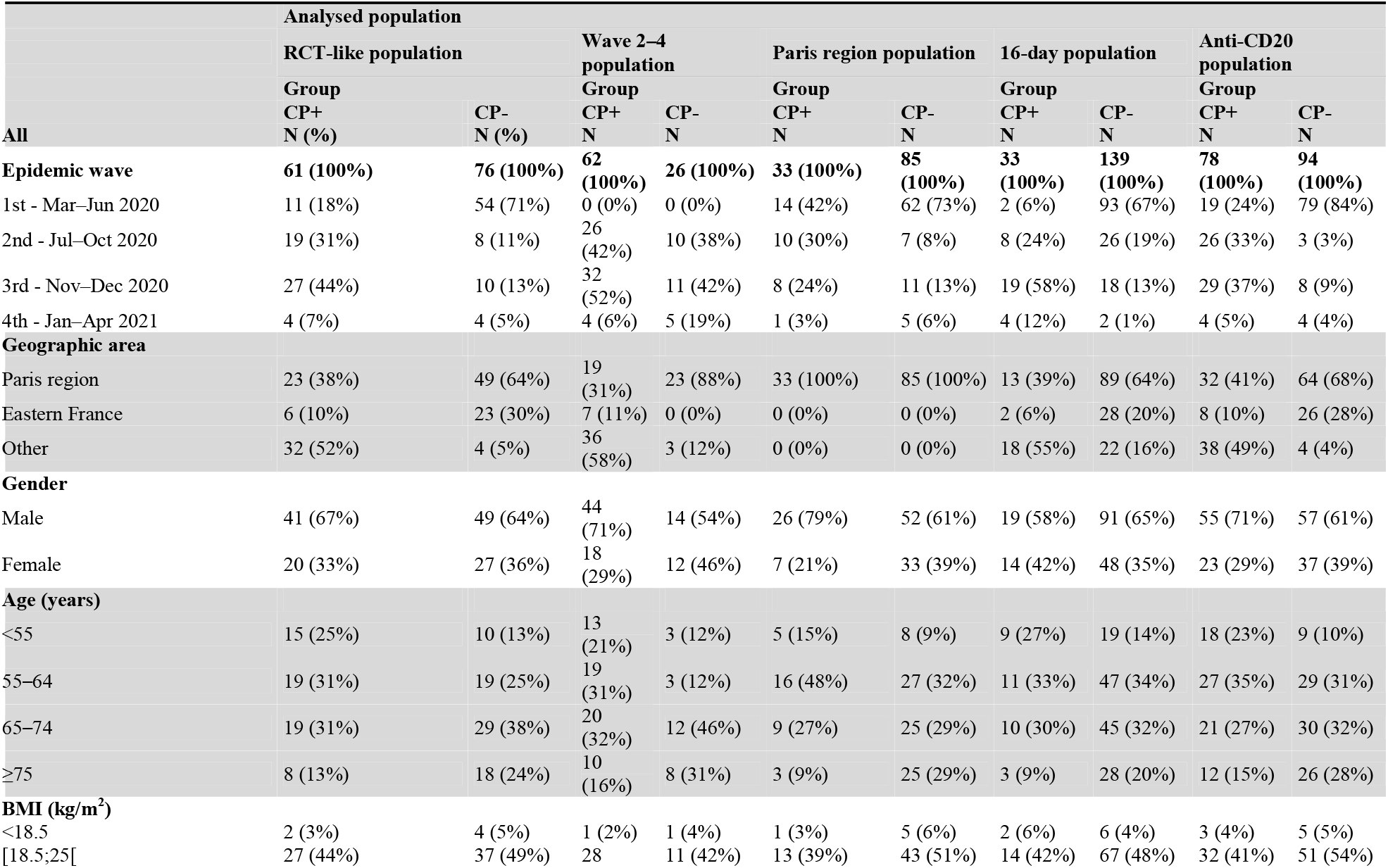
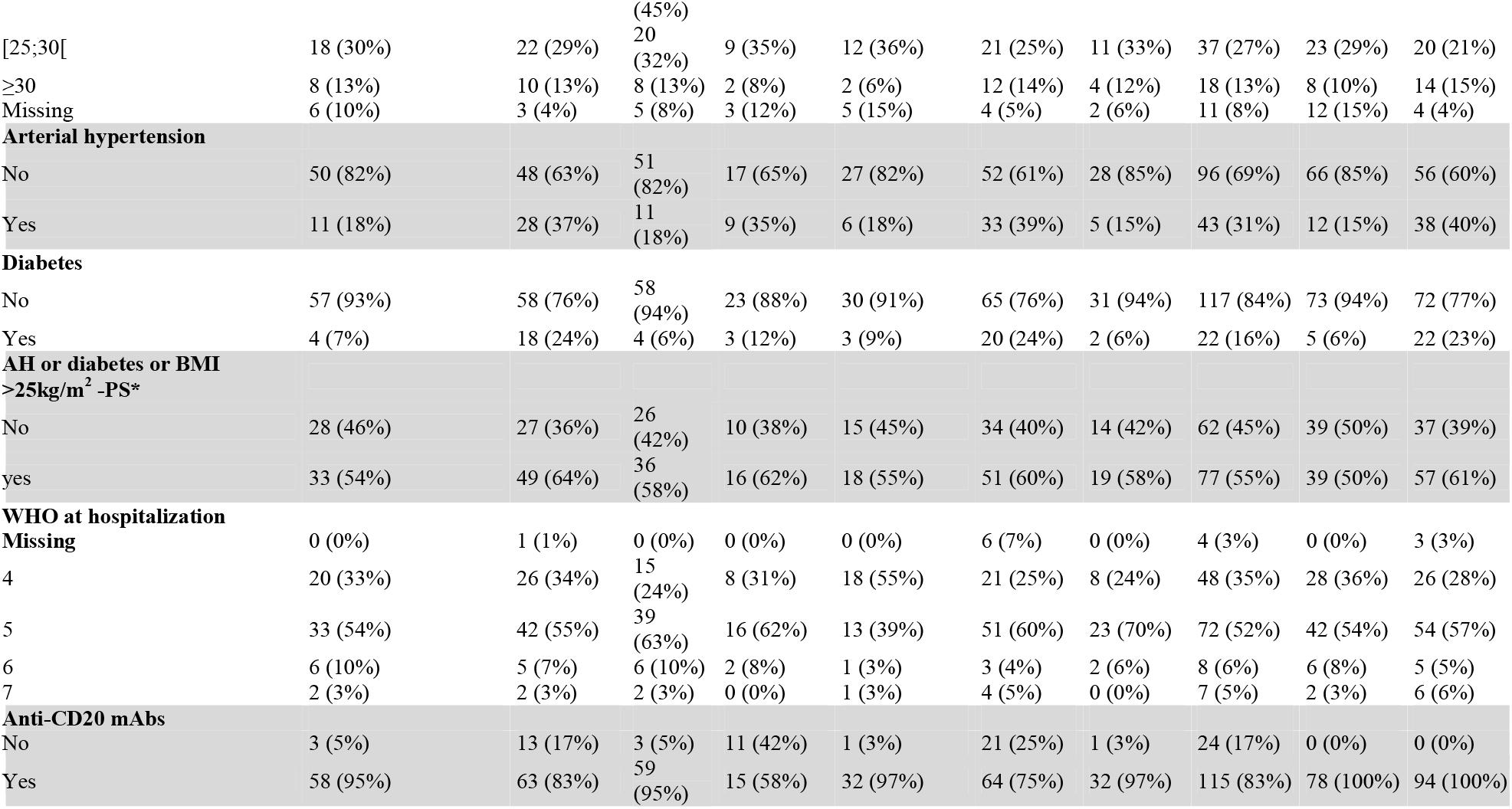
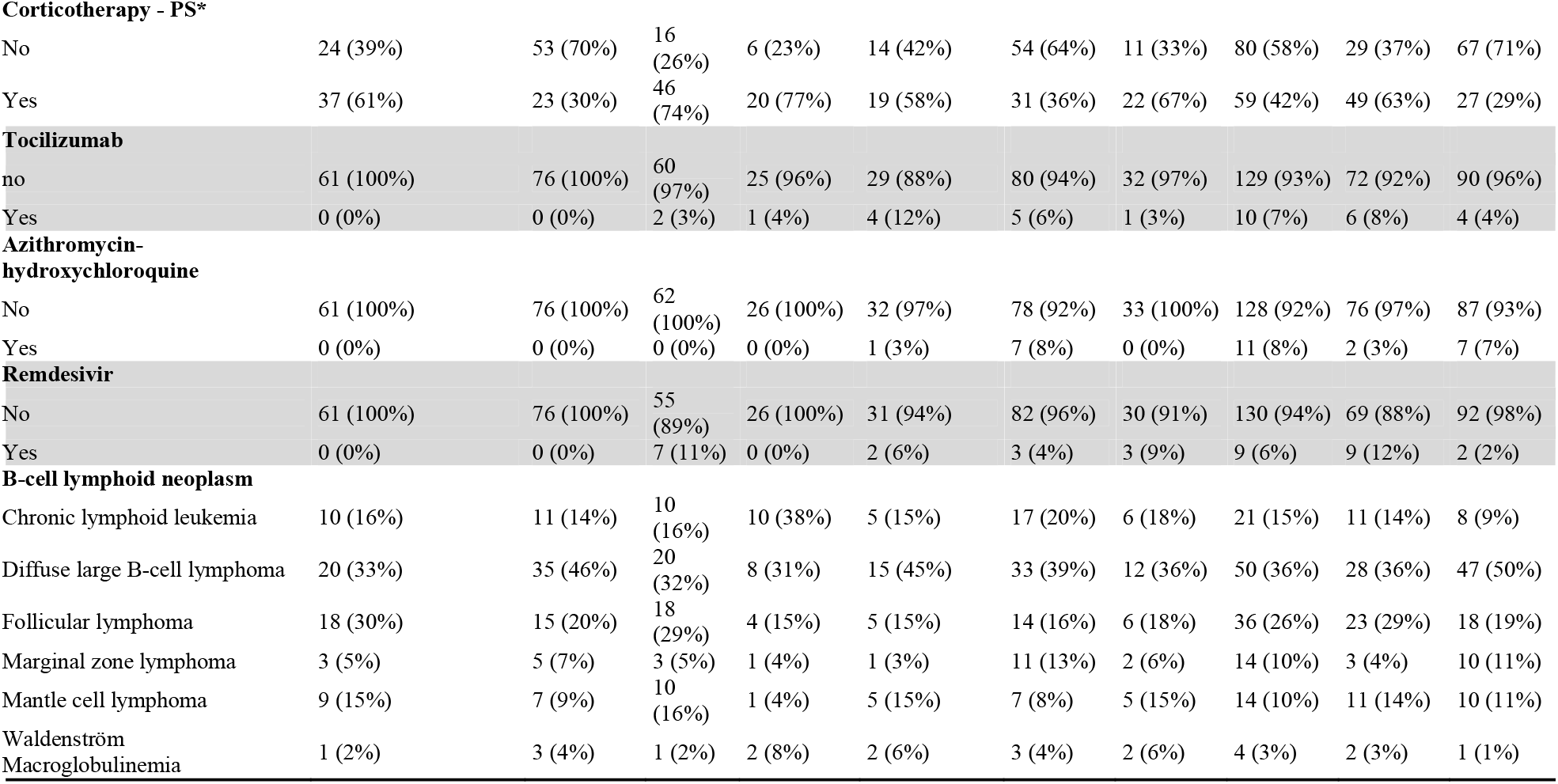
Characteristics of patients according to analyzed population (other sentivity analyses). AH: arterial hypertension, BMI: body mass index, PS*: included in the propensity score, N: number of patients, RCT: randomized control trial, WHO: world health organization, mAbs: monoclonal antibodies

**Table S2.** Estimation of hazard ratio (HR) of death associated with Covid-19 plasma therapy and its 95% confidence interval (CI). IPTW: inverse probability treatment weighting of convalescent plasma exposure, RCT: randomized controlled trial. Confounding factors—age (<65 years vs. ≥65 years), arterial hypertension or diabetes or body mass index >25kg/m2 (yes vs. not), and corticotherapy (yes vs. not)—were included in multivariable Cox proportional models for estimating Adjusted associations* and in weight computation for IPTW** models.

| Analyzed population | N at risk/n deaths | Crude association |  | Adjusted association* |  | IPTW** |  |
| --- | --- | --- | --- | --- | --- | --- | --- |
|  |  | HR [95%CI] | p-value | HR [95%CI] | p-value | HR [95%CI] | p-value |
| 16-day anti-CD20 population (Main analysis) | 147/28 | 0.47 [0.16;1.3] | 0.157 | 0.33 [0.11;0.99] | 0.049 | 0.37 [0.20;0.69] | 0.002 |
| Overall population | 201/64 | 0.45 [0.26;0.79] | 0.005 | 0.46 [0.26;0.84] | 0.012 | 0.50 [0.34;0.72] | <0.001 |
| RCT-like population | 137/42 | 0.54 [0.29;1.0] | 0.064 | 0.58 [0.29;1.2] | 0.117 | 0.57 [0.37;0.89] | 0.014 |
| Wave 2–4 population | 88/26 | 0.44 [0.20;0.97] | 0.042 | 0.50 [0.23;1.1] | 0.083 | 0.60 [0.35;1.0] | 0.069 |
| Paris region population | 118/37 | 0.40 [0.17;0.95] | 0.039 | 0.39 [0.16;0.96] | 0.040 | 0.40 [0.24;0.67] | <0.001 |
| 16-day population | 172/29 | 0.55 [0.19;1.6] | 0.263 | 0.46 [0.15;1.4] | 0.165 | 0.43 [0.23;0.78] | <0.001 |
| Anti-CD20 population | 172/59 | 0.37 [0.21;0.65] | <0.001 | 0.32 [0.17;0.60] | <0.001 | 0.39 [0.26;0.58] | <0.001 |

**Table S3.** Estimation of hazard ratios (HRs) of death associated with covariables included in the multivariable model in the overall population and their 95% confidence intervals (CI). AH: arterial hypertension; BMI: body mass index

| Variable-modality | Adjusted association |  | Adjusted association including sex |  |
| --- | --- | --- | --- | --- |
|  | HR of death<br>[95%CI] | p-value | HR of death<br>[95%CI] | p-value |
| AH or diabetes or BMI >25kg/m<sup>2</sup>: no vs. yes | 0.72 [0.43;1.2] | 0.210 | 0.74 [0.44;1.3] | 0.254 |
| Age (years): <65 vs. ≥65 | 0.50 [0.29;0.85] | 0.011 | 0.50 [0.30;0.86] | 0.012 |
| Corticotherapy: no vs. yes | 0.80 [0.47;1.4] | 0.418 | 0.80 [0.47;1.4] | 0.399 |
| Gender: female vs. male |  |  | 0.82 [0.48;1.4] | 0.481 |
| Exposure to convalescent plasma: yes vs. no | 0.46 [0.26;0.84] | 0.012 | 0.45 [0.25;0.83] | 0.010 |

## Supplementary data – Figures

**Figure S1:**
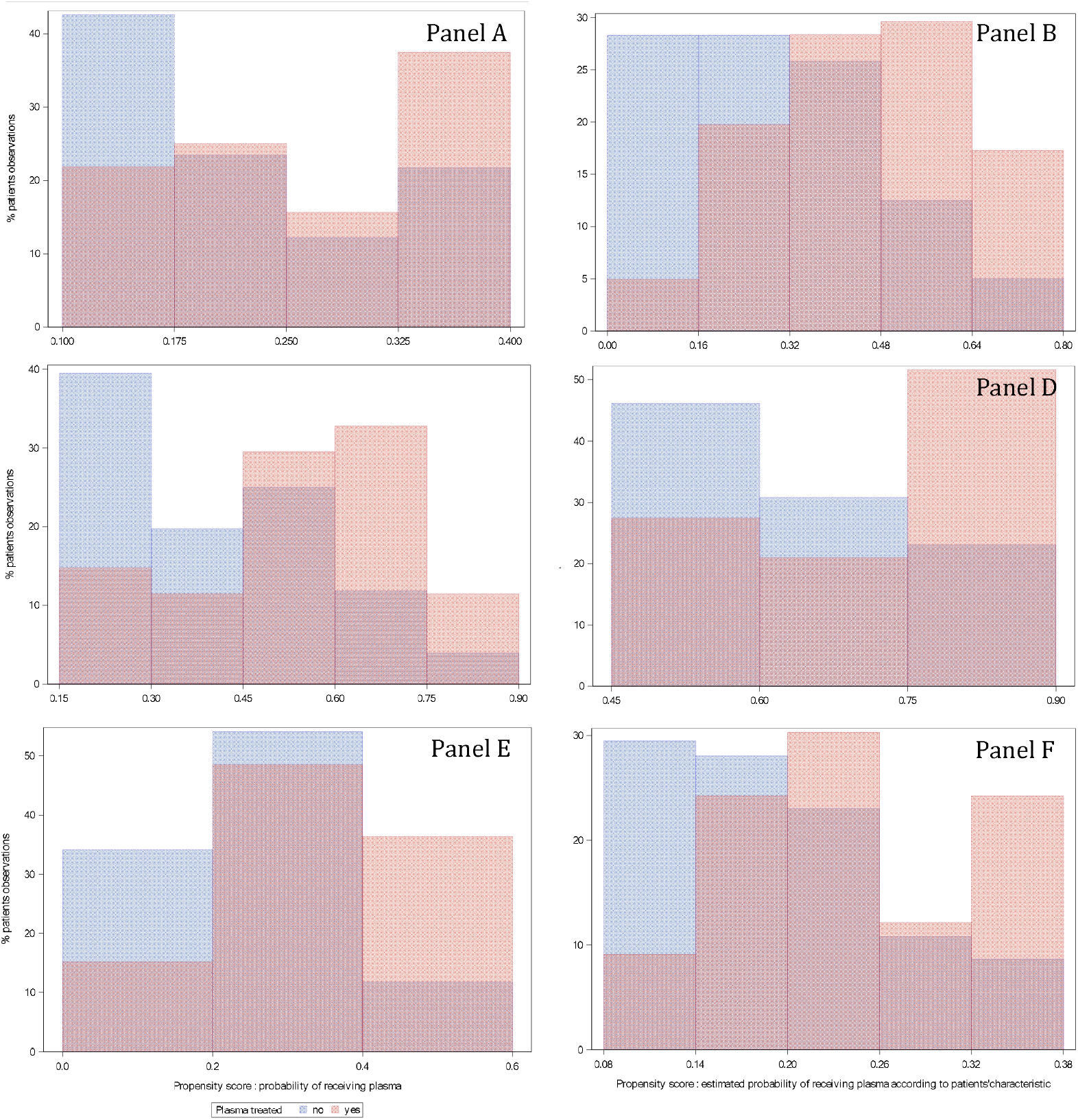
Overlap between the two distributions of propensity scores according to actual exposure to convalescent plasma allowed the use of the propensity score-based method. Distributions of propensity score in the two groups in the 16-day anti-CD20 population (Panel A), in overall population (Panel B), in a randomized controlled trial-like population (Panel C), in wave 2–4 population (Panel D), in the Paris region population (Panel E), and in the 16-day population (Panel F).

**Figure S2:**
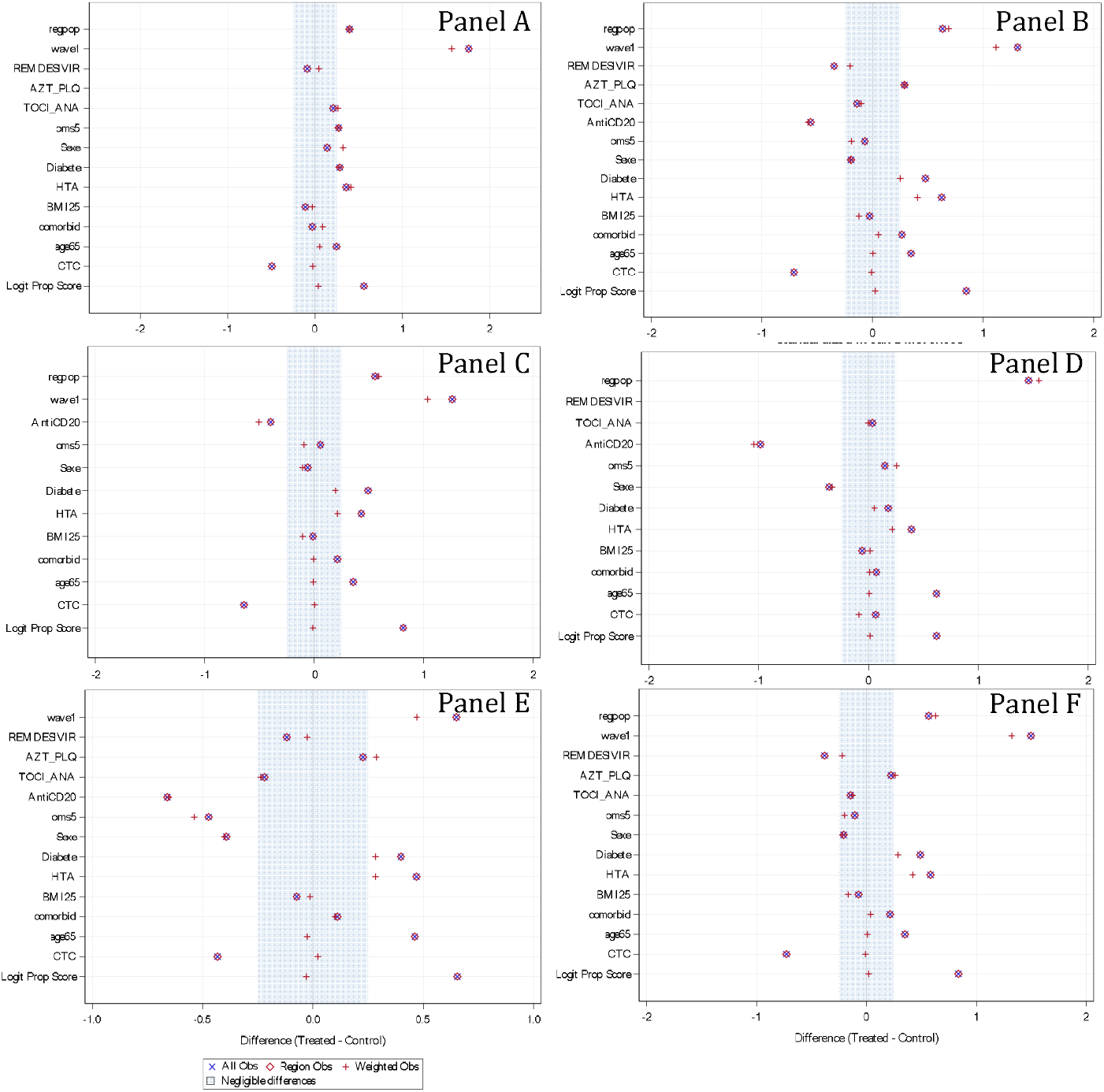
The use of weighted observations succeeded in decreasing the differences between the two groups for variables included in the propensity score. Differences between the two groups could remain for variables that were not included in the propensity score since a very small number of patients could not fit the positivity assumption, and sensitivity analyses were run excluding modalities from these variables. Standardized mean differences between original observations and weighted observations from COVID-19 convalescent plasma (CCP)+ and CCP-groups in 16-day anti-CD20 pre-exposed population (Panel A), in overall population (Panel B), in randomized controlled trial-like population (Panel C), in the wave 2– 4 population (Panel D), in the Paris region population (Panel E), and in the 16-day population (Panel F) for binary variable: wave1: first wave vs. following waves; remdesivir: yes vs. no; AZT_PLQ: azithromycin/hydroxychloroquine yes vs. no, TOCI_ANA: Tocilizumab or Anakinra yes vs. no; OMS 5: WHO score 5 vs. <5; Sexe: male vs. female; Diabete: diabetes yes vs. no; HTA: arterial hypertension

